# Electronic Health Record Coding of Self-Reported Diagnoses in the All of Us Research Program

**DOI:** 10.64898/2026.10.18.26365863

**Authors:** Amir Asiaee, Kaveh Aryan

## Abstract

**Importance:** Conditions that are hard to diagnose, several more common in women, may be missing from electronic health records (EHRs), hiding patients from research cohorts and case finding.

**Objective:** To measure how often the EHR contains a diagnosis code for chronic conditions that participants report, and to compare this with national survey benchmarks.

**Design, Setting, and Participants:** This cross-sectional study used the All of Us Research Program Registered Tier Dataset v9. Participants were adults with EHR visits in both 12-month periods from July 2021 to June 2023 who completed a health history survey. We analyzed the data in October 2026.

**Exposures:** Self-reported clinician diagnosis of 10 conditions with ongoing visits or treatment, as well as sex at birth, race and ethnicity, age, and years of EHR records.

**Main Outcomes and Measures:** Share of participants reporting current care who had a qualifying diagnosis code, alone and relative to hypertension and type 2 diabetes, and the ratio of age-standardized coded shares to national survey prevalence.

**Results:** Of 161,480 respondents, 114,610 (73,870 female [64.5%], median [IQR] age 63 [50–72] years) had at least 5 years of EHR records before the survey. Among those reporting current care, 84.6% (95% CI, 84.2%-85.0%) with hypertension and 87.0% (95% CI, 86.5%-87.6%) with type 2 diabetes had a code, compared with 41.3% (95% CI, 38.9%-43.8%) for endometriosis, 39.4% (95% CI, 34.3%-44.8%) for Lyme disease, and 24.4% (95% CI, 23.2%-25.6%) for chronic fatigue. Counting ICD-9-CM and other mapped codes, 55.3%, 51.5%, and 28.3%, respectively, had any qualifying record. Women had lower coded shares than men for hypertension, type 2 diabetes, obesity, depression, and anxiety (crude ratios 0.93-0.96, within-health-system ratios 0.98-1.02) and higher shares for migraine and fibromyalgia (within health systems, 1.11 [95% CI, 1.07-1.15] and 1.41 [95% CI, 1.29-1.57]). Age-standardized coded shares among active patients were 1.1 to 2.4 times national prevalence of the same self-reported diagnoses.

**Conclusions and Relevance:** In this cross-sectional study, fewer than half of participants reporting ongoing care for chronic fatigue, Lyme disease, or endometriosis had a qualifying ICD-10-CM code. Linking each participant’s report to their own record measured this gap directly.

**Key Points:** *Question:* Among adults who report a clinician-diagnosed chronic condition, how often does their electronic health record contain a corresponding diagnosis code?

*Findings:* In this cross-sectional study of 114,610 All of Us participants with at least 5 years of preceding electronic health records, 83% to 90% of those reporting ongoing care for hypertension or type 2 diabetes had a corresponding code, compared with 41% for endometriosis, 39% for Lyme disease, and 24% for chronic fatigue.

*Meaning:* These findings suggest that cohorts defined by diagnosis codes may omit many patients who report care for chronic fatigue, Lyme disease, or endometriosis.

## Introduction

Diagnosis codes define who is counted in EHR research, which patients appear in quality measures, and which patients decision support can find. A condition that clinicians diagnose but do not code, or that patients are told they have outside the coding health system, can be missed by all three uses. Long COVID shows the problem: its ICD-10-CM code, U09.9, is recorded inconsistently across health systems, with differences in diagnosis by sex, race and ethnicity, insurance, and level of care [1–6]. Similar concerns apply to myalgic encephalomyelitis/chronic fatigue syndrome (ME/CFS) [7, 8], fibromyalgia [9, 10], endometriosis [11], and Lyme disease [12], conditions whose diagnosis is often delayed or disputed and several of which are more common in women.

Two approaches can measure coding without chart review. The first compares the share of a cohort that is coded with survey prevalence in the matching population [13]. It requires the EHR cohort and the survey population to have the same prevalence, which a population defined by contact with care may not. The second links each person’s report of a diagnosis to their own record. Earlier work found substantial agreement between self-report and abstracted records for hypertension and diabetes [14], and a Veterans Health Administration study compared EHR diagnosis rates with survey prevalence across sociodemographic groups for substance use disorders [15]. We extend direct linkage to 10 chronic conditions within a single cohort and compare it with national-survey benchmarks.

In this study, we used the All of Us Research Program, which links survey answers and EHR data for the same participants [16], to measure the share of self-reported diagnoses that carry a code, compare women and men and racial and ethnic groups, and test whether national survey benchmarks give the same answer.

## Methods

### Data and population

We used the All of Us Research Program Registered Tier Dataset v9 (curated data repository R2025Q4R6, with a data cutoff of January 1, 2025), in which dates are shifted backward by 1 to 365 days per participant, preserving intervals within a person. The population was adults born in 2004 or earlier with at least 1 EHR visit in each of 2 consecutive 12-month periods (July 2021 to June 2022 and July 2022 to June 2023), so that each had current contact with a contributing health system. Survey respondents were those who completed the Personal and Family Health History survey or its predecessor, the Personal Medical History survey. The primary analytic population comprised respondents whose first EHR condition record preceded the survey by at least 5 years, so that diagnoses made before the record began were less likely (eTable 1 in the Supplement). Sex was the participant-reported sex at birth in the program record. We analyzed the female and male categories and excluded 2,300 active participants with other or unknown values. We refer to these groups as female and male participants or women and men. Participants self-reported race and ethnicity in the program’s survey. We analyzed these variables because differences in diagnosis by race and ethnicity have been reported for Long COVID [1, 4]. The categories were Hispanic or Latino, non-Hispanic Asian, non-Hispanic Black, non-Hispanic White, non-Hispanic other or multiple races (any other single race or more than 1 race), and unknown. All of Us participants provide informed consent at enrollment under a protocol overseen by the program’s institutional review board. We analyzed de-identified Registered Tier data as authorized researchers [16]. This study followed the Strengthening the Reporting of Observational Studies in Epidemiology (STROBE) reporting guideline [17].

### Conditions and codes

Ten conditions have both a survey item and an ICD-10-CM code: hypertension (I10 to I13, I15, I16), type 2 diabetes (E11), obesity (E66, Z68.3, Z68.4), depression (F32, F33), anxiety (F41), migraine (G43), fibromyalgia (M79.7), endometriosis (N80, among women), Lyme disease (A69.2), and chronic fatigue (G93.3, which includes ME/CFS, or R53.82). We identified the specified ICD-10-CM source codes in the condition table. For obesity, we also searched the observation table. We considered a condition coded if a qualifying code occurred anywhere in the available record. To assess the coverage of this search, we also counted current self-reporters whose only record of the condition was an ICD-9-CM code or a code from another source vocabulary mapped to the same standard concept. For the national comparison, we represented ME/CFS by G93.3 codes alone (ME/CFS and postviral fatigue). Long COVID (U09.9) has no survey item, and we studied it by survey comparison only.

### Self-reported diagnosis

We identified self-reported diagnoses from condition-specific follow-up questions: participants who report a condition for themselves are asked “About how old were you when you were first told you had” it, and any answer to that question defined an ever-told diagnosis (question identifiers and routing checks, eTables 2 and 3 in the Supplement). The current-care definition additionally required answering yes to “Are you still seeing a doctor or health care provider for” the condition or “Are you currently prescribed medications and/or receiving treatment for” it. This definition did not establish that the disease was currently present. We combined answers across survey occasions and counted codes from any date, so the report and the code need not refer to the same period. A sensitivity analysis used only the earliest survey occasion and codes dated no later than 1 year after it (eTable 4 in the Supplement).

### Measures

The primary measure was the share of participants reporting a current diagnosis who had any code for it. Even for routinely coded conditions, this share falls below 1 when care occurs outside the health systems that contribute records or before the record begins. We therefore also report each condition’s share relative to the mean share for 2 reference (anchor) conditions, hypertension and type 2 diabetes, in the same group, requiring both anchor estimates. For endometriosis, the reference was the female anchors. The relative share compares coding with the 2 reference conditions. Interpreting it as condition-specific coding requires shared record incompleteness and complete coding of the reference conditions. For health systems, we assigned each patient to the EHR site that contributed most of their visits from July 2021 to June 2023 (qualifying codes could come from any site). For each condition, we report percentiles across sites of the share coded and of the share relative to that site’s anchors, without identifying sites. A site entered a condition’s percentiles when it had at least 50 current self-reporters of the condition (and of each anchor, for the relative share), so that each site’s share was reasonably precise, and we report percentiles only when at least 5 sites qualified. We also report the women-to-men ratio of the share coded.

We used an operational definition of female predominance: a female-to-male prevalence ratio of at least 1.5 in the selected benchmark (NHIS 2023 or NHANES 2021-2023) or, for fibromyalgia and Lyme disease, in All of Us self-report. The migraine and chronic fatigue classifications used related survey constructs (eTable 5 in the Supplement).

### Survey comparison

For comparison with the cohort-to-survey approach, we computed each condition’s coded share among all active EHR patients, directly standardized within sex to the age distribution (and, separately, the age by race and ethnicity distribution) of US adults from NHANES 2021-2023 interview weights, and divided it by national prevalence for the same sex: measured hypertension or treatment, measured obesity, and diagnosed diabetes from NHANES 2021-2023, and ever-told hypertension, type 2 diabetes, depression, and anxiety, current ME/CFS [8], and ever and current Long COVID from the NHIS 2023 Sample Adult file. For the NHANES obesity and diabetes benchmarks (adults 20 years or older), we computed coded shares among patients of those ages, excluded women with a recent pregnancy code from the obesity comparison, and counted codes for any type of diabetes (eMethods in the Supplement). We computed design-based (Taylor linearization) intervals for survey prevalence [18, 19]. Our NHIS estimates reproduced published chronic pain prevalence [20], and we computed the ME/CFS estimate from NHIS 2023 rather than taking it from the 2021-2022 report [8]. Comparisons between nonequivalent definitions (for chronic pain, migraine, and ME/CFS) are exploratory and are not coding fractions.

### Statistical Analysis

We ran analyses of participant data inside the Researcher Workbench and exported only aggregate tables. We suppressed counts of 1 to 20, rounded other counts up to a multiple of 10, and, within each table, suppressed a further count wherever a suppressed count could otherwise be recovered from the table’s totals (complementary suppression). We calculated shares from the released counts, with 95% Wilson CIs. A sensitivity analysis recomputed them from 1,000 sets of counts compatible with the rounding (eTable 6 in the Supplement). CIs for anchor-relative shares and women-to-men ratios came from a participant-level bootstrap on unrounded data within the Workbench (2,000 replicates, each participant keeping their membership in every condition), with a version resampling health systems (eTable 7 in the Supplement). We also standardized women-to-men ratios for age and record length and estimated them within health systems with the Mantel-Haenszel method (eMethods in the Supplement). We computed standardized shares and site quantiles in the Workbench. Subgroup comparisons were exploratory, and we did not adjust CIs for multiple comparisons. Analyses used Python 3.12 with NumPy 2.4, pandas 2.3, and scikit-learn 1.8 in the Researcher Workbench and, for the released tables, Python 3.10 with pandas 1.5, NumPy 1.26, and matplotlib 3.10.

## Results

### Population

Of 164,840 female and 95,660 male participants with active EHR records, 104,250 (63%) and 57,230 (60%) completed the survey (Table 1). Men were older than women (36% vs 23% aged 70 years or older). Of the 161,480 respondents, 114,610 had at least 5 years of records before the survey (73,870 female, median [IQR] age 63 [50–72] years, 74.6% non-Hispanic White, eTable 1 in the Supplement), and 13,240 of them (11.6%) answered on 2 or more survey occasions.

**Table 1.** Characteristics of Active Adult EHR Patients and the Primary Analytic Population. Active EHR patients: participants aged 18 years or older with at least 1 EHR visit in each of 2 consecutive 12-month periods (July 2021 to June 2023) whose sex at birth was recorded as female or male. Survey respondents completed the Personal and Family Health History (or Personal Medical History) survey. Primary analytic population: survey respondents whose first EHR condition record preceded the survey by 5 years or more (eTable 1 in the Supplement). Age is 2023 minus birth year. We rounded each count up to a multiple of 10 separately, so subgroup counts may not sum to the displayed totals.

| Characteristic | Active EHR patients |  | Primary analytic population |  |
| --- | --- | --- | --- | --- |
|  | Women | Men | Women | Men |
| No. | 164,840 | 95,660 | 73,870 | 40,750 |
| Survey respondents, No. (%) | 104,250 (63) | 57,230 (60) | – | – |
| Age, median (IQR), y | – | – | 60 (46–70) | 68 (57–75) |
| *Age, y, No. (%)* |  |  |  |  |
| 18–39 | 32,850 (20) | 11,840 (12) | 10,880 (15) | 2,850 (7) |
| 40–49 | 25,930 (16) | 10,630 (11) | 11,090 (15) | 3,560 (9) |
| 50–59 | 30,820 (19) | 14,850 (16) | 13,910 (19) | 5,640 (14) |
| 60–69 | 37,990 (23) | 24,250 (25) | 18,390 (25) | 10,600 (26) |
| 70 or older | 37,260 (23) | 34,100 (36) | 19,630 (27) | 18,120 (44) |
| *Race and ethnicity, No. (%)* |  |  |  |  |
| Hispanic or Latino | 26,090 (16) | 11,820 (12) | 7,030 (10) | 2,970 (7) |
| Non-Hispanic Asian | 4,710 (3) | 3,000 (3) | 1,450 (2) | 910 (2) |
| Non-Hispanic Black | 23,660 (14) | 11,410 (12) | 6,870 (9) | 2,440 (6) |
| Non-Hispanic White | 98,400 (60) | 62,140 (65) | 53,700 (73) | 31,820 (78) |
| Non-Hispanic other or multiple | 8,120 (5) | 4,500 (5) | 3,250 (4) | 1,560 (4) |
| Unknown | 3,880 (2) | 2,800 (3) | 1,600 (2) | 1,060 (3) |

### Share of self-reported diagnoses that were coded

Among participants with at least 5 years of records who reported current care, the anchors were coded for 83% (women) and 87% (men) with hypertension and 84% and 90% with type 2 diabetes (Table 2, with all record lengths in Figure 1A and eTable 8 in the Supplement). Relative to the anchors, the share coded was 0.88 (95% CI, 0.87-0.89) for obesity and 0.78 to 0.79 for depression, anxiety, and migraine. It was 0.73 (95% CI, 0.71-0.75) for fibromyalgia, 0.49 (95% CI, 0.46-0.52) for endometriosis, 0.46 (95% CI, 0.39-0.51) for Lyme disease, and 0.28 (95% CI, 0.27-0.30) for chronic fatigue. Using only the earliest survey occasion changed the shares by less than 0.01, and also limiting codes to the year after it lowered them by 0.05 or less (eTable 4 in the Supplement). Six of the 7 conditions below obesity were female predominant. Shares were lower for ever-told diagnoses, most for conditions often diagnosed long ago: among women aged 70 years or older, 6% of those ever told they had endometriosis had a code.

**Figure 1.**
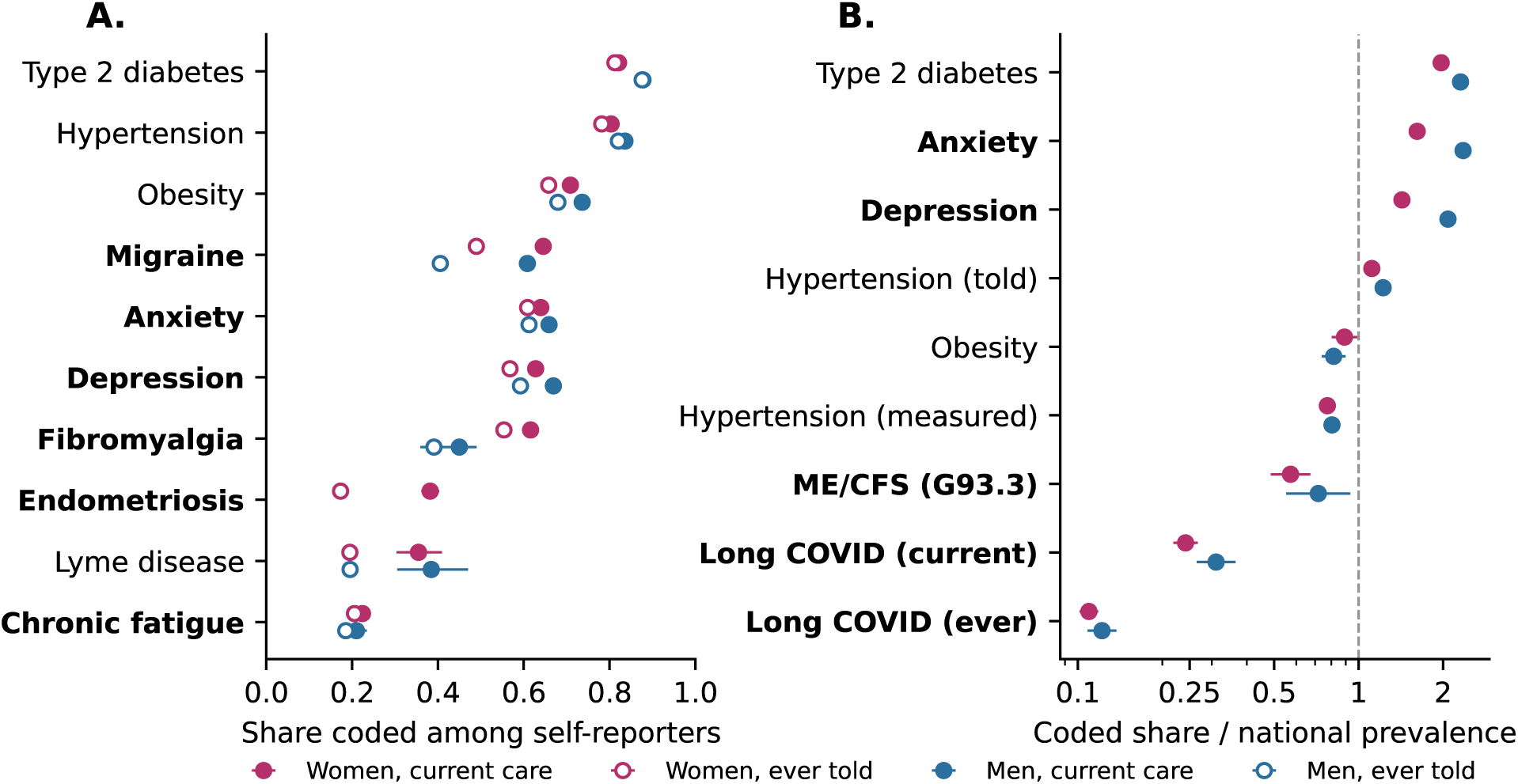
Coding of Self-Reported Diagnoses and Comparison With National Prevalence. **A.** Share with any EHR code among survey respondents who report the diagnosis, by sex: currently treated or still seeing a clinician (filled) or ever told (open), all lengths of EHR record. **B.** Coded share among all active EHR patients, standardized within sex to the age distribution of US adults, divided by national prevalence for the same sex (log scale, with the dashed line at equality): hypertension (measured or treated from NHANES 2021-2023, ever told from NHIS 2023), obesity (measured, NHANES, adults 20 years or older), and type 2 diabetes, depression, anxiety, ME/CFS (current, with ME/CFS and postviral fatigue codes [G93.3] in the numerator), and Long COVID (ever and current) from NHIS 2023. Bold labels mark female-predominant conditions. Error bars are 95% CIs.

**Table 2.** Share With a Qualifying Code Among Participants Reporting Current Care. Survey respondents with 5 or more years of EHR condition records before the survey who reported a clinician diagnosis and ongoing visits or treatment for the condition. No.: respondents reporting current care (rounded up to a multiple of 10). Shares are proportions with 95% Wilson CIs from the released counts. The women-to-men ratio and the share relative to anchors have participant-level bootstrap 95% CIs (eMethods in the Supplement). Relative to anchors: the share divided by the mean share for hypertension and type 2 diabetes among women and men pooled (for endometriosis, the female anchors). Bold: female predominant (eTable 5 in the Supplement).

| Condition | No. | All | Women | Men | Women-to-men ratio | Relative to anchors<sup>a</sup> |
| --- | --- | --- | --- | --- | --- | --- |
| Hypertension | 37,360 | 0.85 (0.84–0.85) | 0.83 (0.83–0.84) | 0.87 (0.86–0.87) | 0.96 (0.95–0.97) | Reference |
| Type 2 diabetes | 13,350 | 0.87 (0.86–0.88) | 0.84 (0.83–0.85) | 0.90 (0.89–0.91) | 0.94 (0.92–0.95) | Reference |
| Obesity | 16,080 | 0.75 (0.75–0.76) | 0.74 (0.74–0.75) | 0.78 (0.77–0.79) | 0.95 (0.93–0.97) | 0.88 (0.87–0.89) |
| **Depression** | 28,130 | 0.68 (0.67–0.68) | 0.67 (0.66–0.67) | 0.71 (0.70–0.72) | 0.93 (0.92–0.95) | 0.79 (0.78–0.80) |
| **Anxiety** | 21,450 | 0.68 (0.67–0.69) | 0.67 (0.67–0.68) | 0.70 (0.69–0.72) | 0.96 (0.94–0.98) | 0.79 (0.78–0.80) |
| **Migraine** | 12,970 | 0.67 (0.66–0.68) | 0.68 (0.67–0.69) | 0.64 (0.62–0.66) | 1.06 (1.03–1.10) | 0.78 (0.77–0.79) |
| **Fibromyalgia** | 5,350 | 0.63 (0.61–0.64) | 0.64 (0.63–0.65) | 0.48 (0.43–0.52) | 1.34 (1.24–1.51) | 0.73 (0.71–0.75) |
| **Endometriosis** | 1,500 | 0.41 (0.39–0.44) | 0.41 (0.39–0.44) | NA | NA | 0.49 (0.46–0.52) |
| Lyme disease | 330 | 0.39 (0.34–0.45) | 0.39 (0.33–0.46) | 0.40 (0.31–0.50) | 0.98 (0.73–1.32) | 0.46 (0.39–0.51) |
| **Chronic fatigue** | 4,880 | 0.24 (0.23–0.26) | 0.25 (0.23–0.26) | 0.23 (0.20–0.26) | 1.09 (0.95–1.23) | 0.28 (0.27–0.30) |

<sup>a</sup>The anchors' own values relative to their mean are 0.99 (hypertension) and 1.01 (type 2 diabetes). NA, not applicable (endometriosis was analyzed among participants recorded as female at birth).

ICD-9-CM codes and codes from other mapped vocabularies explained part of the shortfall. Counting them, 95.6% of current self-reporters with hypertension and 97.5% with type 2 diabetes had a qualifying record, compared with 55.3% with endometriosis, 51.5% with Lyme disease, and 28.3% with chronic fatigue (eTable 9 in the Supplement).

The anchors’ share coded rose with the length of the record, from 75% (hypertension, under 2 years of records before the survey) to 86% (10 years or more) (Figure 2B and eTable 10 in the Supplement). Shares for the other conditions rose as well, but with 10 years or more of records, 25% of participants reporting current chronic fatigue, 39% reporting current Lyme disease, and 41% reporting current endometriosis had a code.

**Figure 2.**
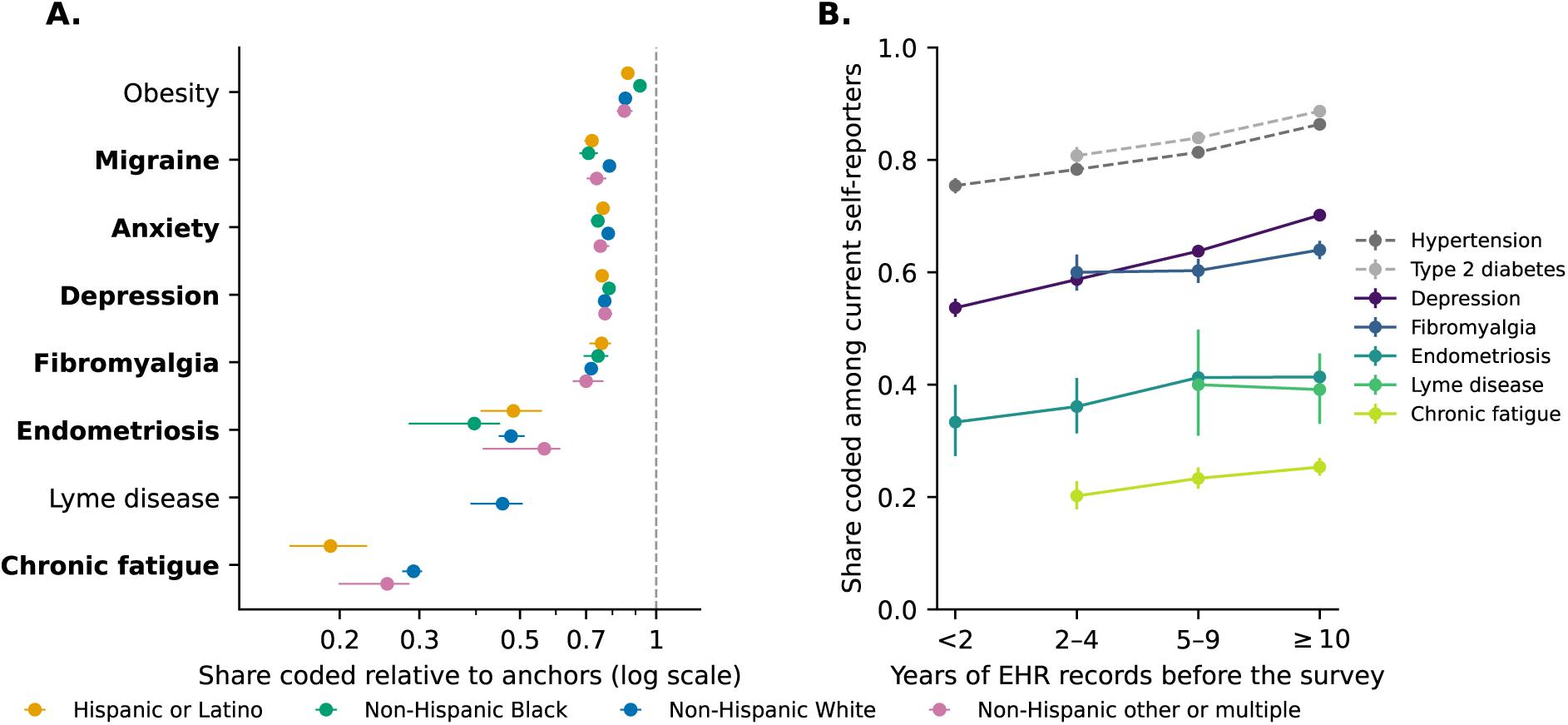
Coding of Current Self-Reported Diagnoses by Race and Ethnicity and by Length of EHR Record. **A.** Share of current self-reporters with a code relative to the mean share for the anchor conditions (hypertension and type 2 diabetes) in the same racial and ethnic group (for endometriosis, the female anchors), all lengths of record (log scale). The dashed line marks equality with the anchors, and error bars are participant-level bootstrap 95% CIs. We do not show groups whose estimates were suppressed for privacy. We do not show groups in which an anchor estimate was suppressed (including non-Hispanic Asian participants). **B.** Share of current self-reporters with a code by years of EHR condition records before the survey. Dashed lines mark the anchor conditions, and error bars are 95% Wilson CIs. We pooled women and men (endometriosis, women only). Bold labels mark female-predominant conditions.

### Health systems

Across the 51 health systems with at least 50 current self-reporters of hypertension, the share with a hypertension code ranged from 0.30 at the 10th percentile to 0.97 at the 90th (median, 0.92). This variation may reflect record completeness, patient composition, and coding practices. Relative to each site’s anchors, shares varied less. For chronic fatigue, the median share relative to the site’s anchors was 0.29 across 30 sites (10th-90th percentile, 0.13-0.43), and for endometriosis, relative to the female anchors, 0.49 across 12 sites (10th-90th percentile, 0.18-0.69). Fewer than 5 sites had enough patients for Lyme disease (eTable 11 in the Supplement).

### Sex and race and ethnicity

In unadjusted comparisons among respondents reporting current care for the same condition, women were coded less often than men for hypertension (ratio, 0.96; 95% CI, 0.95-0.97), type 2 diabetes (ratio, 0.94; 95% CI, 0.92-0.95), obesity (ratio, 0.95; 95% CI, 0.93-0.97), depression (ratio, 0.93; 95% CI, 0.92-0.95), and anxiety (ratio, 0.96; 95% CI, 0.94-0.98), and more often for migraine (ratio, 1.06; 95% CI, 1.03-1.10) and fibromyalgia (ratio, 1.34; 95% CI, 1.24-1.51) (Table 2). Standardized for age and record length, the first 5 ratios were 0.95 to 0.98, and within health systems they were 0.98 to 1.02, whereas migraine (ratio, 1.11; 95% CI, 1.07-1.15) and fibromyalgia (ratio, 1.41; 95% CI, 1.29-1.57) remained higher for women (eTable 12 in the Supplement). Crude ratios for Lyme disease (ratio, 0.98; 95% CI, 0.73-1.32) and chronic fatigue (ratio, 1.09; 95% CI, 0.95-1.23) were imprecise. We cannot attribute these differences to clinician behavior.

Anchor means ranged from 0.80 to 0.86 across racial and ethnic groups. Relative to them (Figure 2A and eTable 13 in the Supplement), chronic fatigue was coded less often for Hispanic than for non-Hispanic White participants (0.19 [95% CI, 0.16-0.23] vs 0.29 [95% CI, 0.28-0.30]) and migraine less often for non-Hispanic Black (0.71 [95% CI, 0.68-0.74]) and Hispanic (0.72 [95% CI, 0.69-0.75]) participants than for non-Hispanic White participants (0.79 [95% CI, 0.78-0.80]). Obesity was coded more often for non-Hispanic Black than for non-Hispanic White participants (0.92 [95% CI, 0.90-0.94] vs 0.85 [95% CI, 0.85-0.86]). Relative to the female anchors, endometriosis was coded 0.48 (95% CI, 0.41-0.56) for Hispanic, 0.40 (95% CI, 0.28-0.45) for non-Hispanic Black, and 0.48 (95% CI, 0.45-0.51) for non-Hispanic White participants, and it fell from 0.65 (95% CI, 0.59-0.70) at ages 18 to 39 years to 0.42 (95% CI, 0.36-0.49) at 50 to 59 years (eTable 14 in the Supplement). Requiring an explicit non-Hispanic response for the non-Hispanic categories changed no displayed estimate.

### Comparison with national surveys

After age standardization, coded shares among active EHR patients exceeded national prevalence of the same self-reported diagnoses for hypertension (1.12 for women and 1.22 for men), depression (1.43 and 2.08), anxiety (1.62 and 2.36), and type 2 diabetes (1.97 and 2.31) (Figure 1B and eTable 15 in the Supplement), and for diagnosed diabetes of any type among adults 20 years or older (1.52 and 1.62). Coded shares were lower than measured prevalence of hypertension (0.77 and 0.80) and, among adults 20 years or older, obesity (0.89 and 0.82), consistent with undiagnosed disease. For Long COVID, the coded share was 0.11 (95% CI, 0.10-0.12) of national prevalence of ever having it among women and 0.12 (95% CI, 0.11-0.14) among men, and 0.24 (95% CI, 0.22-0.27) and 0.31 (95% CI, 0.27-0.36) of current prevalence. For G93.3 codes, the coded share was 0.57 (95% CI, 0.49-0.67) of current ME/CFS prevalence among women and 0.72 (95% CI, 0.55-0.94) among men. Standardizing additionally for race and ethnicity changed these ratios by less than 5%.

## Discussion

In a cohort with current EHR records and linked survey answers, most participants who reported a current diagnosis of chronic fatigue, Lyme disease, or endometriosis had no qualifying ICD-10-CM code for it, while more than 80% of those reporting hypertension or type 2 diabetes did. Fibromyalgia, migraine, and mood disorders fell between. Chronic fatigue, Lyme disease, and endometriosis remained the least coded when ICD-9-CM and other mapped codes were counted and when only the earliest survey was used.

Earlier work found substantial agreement between self-report and abstracted medical records for hypertension and diabetes, although its validation measures differ from the conditional coding shares we estimated [14]. Earlier Lyme disease chart studies found the specific ICD-9 code in 43% of records meeting case definitions [12]. Differences in reference standards and coding systems limit comparison with our estimate. For endometriosis, the difference between ever-reported and current-care estimates may reflect historical diagnoses, changes in care, and incomplete records, in keeping with long intervals from symptoms to diagnosis [11]. We did not assess diagnosis timing relative to record onset.

The crude sex differences ran in 2 directions. Women’s lower coding shares for hypertension, type 2 diabetes, obesity, depression, and anxiety shrank after standardization and were close to 1 within health systems, so they mainly reflect how women and men were distributed across health systems, ages, and record lengths. Survey-based gaps, such as the lower Long COVID coding fraction among women [13], may need the same within-system comparison. For fibromyalgia and migraine, men were coded less often, also within health systems. One reading is that coding follows clinicians’ expectations of who has a condition, which this study cannot test. The fibromyalgia difference may also reflect what self-report measures: many people reporting a clinical diagnosis of fibromyalgia do not meet survey criteria [10], and that share may differ by sex.

The survey comparison shows why a within-cohort reference is useful. For common conditions the coded share exceeded national prevalence, by up to 2.4-fold and by different amounts for different conditions, so a survey ratio cannot rank coding across conditions. Differences between coded shares and national prevalence may reflect population selection, measurement differences, and incomplete coding. These comparisons do not separately identify those mechanisms or establish the coding fractions for Long COVID or ME/CFS.

Cohorts defined solely by the selected codes may omit participants who report care for these conditions. The implications for quality measures, registries, and case-finding tools depend on their eligibility and labeling rules. Models trained exclusively on diagnosis codes may learn patterns specific to coded patients. Long COVID phenotyping studies have also used chart review and other reference standards to address the limitations of code-based labels [21, 22].

A companion study uses these reports as training labels for case-finding models [23], and another describes software that delivers such models to patients and clinicians [24].

### Limitations

Self-report measures a diagnosis that a patient was given, not true disease, and recall and interpretation of the questions may differ by condition and group. The current-care definition is a proxy for care status. Survey answers and codes need not describe the same period, although the earliest-survey analysis changed the shares little. Diagnoses recorded only in free text or outside contributing health systems would appear uncoded. The anchor comparison removes record incompleteness only if it is the same across conditions, which the site analysis does not verify, and the restriction to 5 or more years of records reduces but does not remove diagnoses that predate the record. We did not adjust comparisons by race and ethnicity, and we adjusted sex comparisons only for age, record length, and health system. Wilson intervals do not reflect rounding of released counts, which changed the estimates little (eTable 6 in the Supplement). The survey’s chronic fatigue item does not name ME/CFS, and the code set mixes ME/CFS with unspecified chronic fatigue. About 60% of active patients completed the survey, and All of Us participants differ from the US population [25]. We suppressed some estimates for privacy, mainly Lyme disease estimates for most racial and ethnic groups and for adults younger than 50 years, and endometriosis estimates for women aged 70 years or older.

## Conclusions

In this cross-sectional study, fewer than half of respondents reporting ongoing care for chronic fatigue, Lyme disease, or endometriosis had a qualifying ICD-10-CM code. Linked self-report and EHR data measured this discordance directly. Comparisons with national prevalence addressed a different quantity.

## Data Availability

The All of Us Registered Tier Dataset v9 is available to authorized researchers through the All of Us Researcher Workbench. The NHANES 2021-2023 and NHIS 2023 public-use files are available from the National Center for Health Statistics. The aggregate statistics behind every table and figure, and the code and a notebook that regenerate them, are available at https://github.com/AsiaeeLab/unom-self-report-coding.

## Author Contributions

Dr Asiaee had full access to all of the data in the study and takes responsibility for the integrity of the data and the accuracy of the data analysis. *Concept and design:* Asiaee, Aryan. *Acquisition, analysis, or interpretation of data:* Asiaee. *Drafting of the manuscript:* Asiaee. *Critical review of the manuscript for important intellectual content:* Asiaee, Aryan. *Statistical analysis:* Asiaee. *Administrative, technical, or material support:* Aryan.

## Conflict of Interest Disclosures

None reported.

## Funding/Support

None reported.

## Additional Contributions

We gratefully acknowledge All of Us participants for their contributions, without whom this research would not have been possible. We also thank the National Institutes of Health’s All of Us Research Program for making available the participant data examined in this study. The All of Us Research Program is supported by the National Institutes of Health, Office of the Director: Regional Medical Centers: 1 OT2 OD026549; 1 OT2 OD026554; 1 OT2 OD026557; 1 OT2 OD026556; 1 OT2 OD026550; 1 OT2 OD 026552; 1 OT2 OD026553; 1 OT2 OD026548; 1 OT2 OD026551; 1 OT2 OD026555; IAA #: AOD 16037; Federally Qualified Health Centers: HHSN 263201600085U; Data and Research Center: 5 U2C OD023196; Biobank: 1 U24 OD023121; The Participant Center: U24 OD023176; Participant Technology Systems Center: 1 U24 OD023163; Communications and Engagement: 3 OT2 OD023205; 3 OT2 OD023206; and Community Partners: 1 OT2 OD025277; 3 OT2 OD025315; 1 OT2 OD025337; 1 OT2 OD025276. In addition, the All of Us Research Program would not be possible without the partnership of its participants.

## Data Sharing Statement

This study used data from the All of Us Research Program’s Registered Tier Dataset v9, available to authorized users on the Researcher Workbench. NHANES and NHIS data are public. A public repository (MIT License) contains the analysis code and the aggregate statistics behind every table and figure: https://github.com/AsiaeeLab/unom-self-report-coding.

## Notes

### Competing Interest Statement

The authors have declared no competing interest.

### Author Declarations

All of Us Research Program, National Institutes of Health: participants provide informed consent under the program protocol approved by the All of Us Institutional Review Board. We analyzed de-identified data from the All of Us Registered Tier Dataset v9 as authorized researchers on the All of Us Researcher Workbench (https://www.researchallofus.org).

