## Supplementary Material for "Electronic Health Record Coding of Self-Reported Diagnoses in the All of Us Research Program"

### eMethods

#### Population

The active EHR population comprised All of Us participants in the Registered Tier Dataset v9 (curated data repository R2025Q4R6, with a data cutoff of January 1, 2025) born in 2004 or earlier with at least 1 visit in the EHR visit table in each of 2 consecutive 12-month periods, July 1, 2021, to June 30, 2022, and July 1, 2022, to June 30, 2023 (participant-specific shifted dates). Survey respondents had a survey record for the Personal and Family Health History survey or the earlier Personal Medical History survey. Every participant who answered a follow-up diagnosis question was also in this set. We defined age as 2023 minus year of birth (approximate age in 2023), and eligibility required birth in 2004 or earlier. We grouped race and ethnicity as Hispanic or Latino (any race), non-Hispanic Asian, non-Hispanic Black, non-Hispanic White, non-Hispanic other or multiple races, and unknown (skipped, prefer not to answer, or no matching concept). The program's mapping assigns a non-Hispanic category by race whenever ethnicity is not Hispanic or Latino, without requiring an explicit non-Hispanic response. Among active participants, every participant assigned to the non-Hispanic Asian, Black, or White category had answered Not Hispanic or Latino. The mapping assigned 50 participants with no matching ethnicity concept to non-Hispanic other or multiple races. In a sensitivity analysis, we required an explicit Not Hispanic or Latino response for the non-Hispanic categories and classed these participants as unknown. Every respondent count, current self-report count, and coded count shown for both mappings by race and ethnicity (156 cells), and every age by race and ethnicity standardized coded share, was identical. Eligibility steps and counts are in eTable 1.

#### Self-reported diagnosis

The Personal and Family Health History survey asks, by organ system, "Have you or anyone in your family ever been diagnosed with the following ... conditions? Think only of the people you are related to by blood. Select all that apply," and for each selected condition, "Including yourself, who in your family has had [condition]? Select all that apply." Participants who select themselves are shown follow-up questions, including "About how old were you when you were first told you had [condition]?". Because that question is shown only after a participant selects themselves, any recorded answer to it, including a skipped answer, identified an ever-told diagnosis. The earlier Personal Medical History survey asks the same follow-up questions after a participant selects the condition in a list of their own conditions, which all 70,850 respondents to that survey reached. Lyme disease is asked through a question about the respondent alone, shown to every respondent, rather than through the family-history question. We checked this routing for each condition among all respondents (eTable 3): of participants who selected themselves for a condition, all but at most 20 per condition had a first-told answer, and no more than 20 per condition (none for 6 conditions) had a first-told answer without selecting themselves. Counted within each survey separately, up to 60 per condition selected themselves without a first-told answer in that survey. Skipped first-told answers, counted as ever told, numbered 40 to 200 per condition, and no participant with a first-told answer lacked an answer to both current-care questions. The survey version number was not populated in the data, so we could check routing by survey but not by survey version. A current diagnosis required an ever-told diagnosis and "Yes" to either follow-up question on current care (eTable 2).

We defined a survey occasion as one questionnaire response, linking each answer to its survey record (answers without a questionnaire response identifier formed 1 occasion per answer date). In the primary analytic population, 13,240 of 114,610 participants (11.6%) answered on 2 or more occasions. In the sensitivity analysis (eTable 4), current care had to be reported on the participant's earliest occasion, and a code counted if it was dated no later than 1 year after that occasion (any earlier date counted).

#### EHR record length

Record length was the time from a participant's first EHR condition record (any condition) to their earliest survey completion date. The main analysis used participants with 5 years or more. Every respondent had a survey date. Of the respondents, 2,400 had no EHR condition record, so their record length was undefined, and we omitted them from the record-length results (eTable 10). Another 4,140 had a first condition record after the survey (eTable 1).

#### Coverage of the code search

For current self-reporters in the primary analytic population without a qualifying ICD-10-CM source code, we checked first for a qualifying ICD-10-CM code in the observation table, then for an ICD-9-CM code for the condition (patterns in eTable 9), and last for a code from another source vocabulary (not ICD-10-CM and not a survey vocabulary) recorded with a standard concept to which a qualifying ICD-10-CM code maps one to one. R53.82 has no ICD-9-CM equivalent. We counted each participant in the first category that applied.

#### Anchor-relative share

For condition $k$ in group $g$, let $s_{kg}$ be the share of current self-reporters with a code. The relative share is $r_{kg} = s_{kg}/\bar{s}_{Ag}$, where $\bar{s}_{Ag}$ is the mean of the shares for hypertension and type 2 diabetes in the same group. If care outside contributing health systems, or before the record begins, reduces the share coded by the same factor $\lambda_g$ for every condition, so that $s_{kg} = \lambda_g c_{kg}$ with $c_{kg}$ the condition-specific coding probability, then $r_{kg} = c_{kg}/\bar{c}_{Ag}$, which equals $c_{kg}$ when the anchors are coded whenever they are managed. For endometriosis, $\bar{s}_{Ag}$ is the mean of the anchor shares among women in the same group. We obtained intervals for $r_{kg}$ from a participant-level bootstrap inside the Researcher Workbench. In each of 2,000 replicates, every respondent received a Poisson(1) weight that applied to all of their condition memberships, and we recomputed $r_{kg}$ from the weighted, unrounded counts. The 2.5th and 97.5th percentiles formed the interval. Because the same participants enter the condition's share and both anchor shares, the interval includes the uncertainty in $\bar{s}_{Ag}$ and the overlap between conditions. A site-cluster version resampled assigned EHR sites (participants without an assigned site formed 1 cluster). We exported only interval endpoints, and point estimates come from the released counts. For the primary estimates, the earlier approximation (the Wilson limits for $s_{kg}$ divided by a fixed $\bar{s}_{Ag}$) had nearly the same width as the participant bootstrap, and the site-cluster intervals were 2 to 10 times as wide (eTable 7).

#### Women-to-men ratios

The crude ratio is the women's share divided by the men's share (Table 2 in the main text), with a participant-level bootstrap interval as above. The standardized ratio directly standardized each sex's share to the pooled distribution of current self-reporters of both sexes over 10 strata (5 age bands by 2 record-length groups, 5 to 9 years and 10 years or more), using strata with both sexes. The within-site ratio was the Mantel-Haenszel risk ratio over strata of assigned EHR site, age band, and record length. Both used the bootstrap for intervals (eTable 12).

#### Survey comparison

Within each sex, we directly standardized the coded share among all active EHR patients as $\sum_j w_j \hat{p}_j$, where $\hat{p}_j$ is the coded share in stratum $j$ (age band, or age band by race and ethnicity, excluding unknown race and ethnicity) and $w_j$ is the share of US adults of that sex in stratum $j$ (NHANES August 2021-August 2023 demographic file, interview weights, adults 18 years or older). Its standard error was $\{\sum_j w_j^2 \hat{p}_j(1-\hat{p}_j)/n_j\}^{1/2}$. The ratio to national prevalence $\pi$ had a log-scale interval combining both standard errors. For the NHANES obesity and diagnosed-diabetes benchmarks, which cover adults 20 years or older, we computed the All of Us share among active patients aged 20 years or older and standardized it with weights for US adults aged 20 years or older. For obesity among women, we excluded 6,760 women aged 20 years or older with an ICD-10-CM pregnancy code (O00-O9A, Z33, Z34, or Z3A) dated July 2021 to June 2023, as NHANES excludes pregnant women. Without this exclusion, the ratio was 0.91. Because NHANES asks about diagnosed diabetes of any type, that comparison used codes E08, E09, E10, E11, and E13. The NHIS type 2 diabetes comparison used E11.

#### National benchmarks

In NHANES August 2021-August 2023, we defined hypertension as mean systolic blood pressure 130 mm Hg or higher, mean diastolic 80 mm Hg or higher (up to 3 oscillometric readings), or current medication. We defined obesity as measured body mass index of 30 or higher (pregnant women excluded) and diagnosed diabetes as a self-reported diagnosis of any type. From the NHIS 2023 Sample Adult file, we used ever told hypertension, diabetes (type 2 from the type question), depression, and anxiety, as well as ME/CFS ever told and still present. We defined Long COVID as symptoms lasting 3 months or longer after COVID-19 (ever) and symptoms now (current), among all adults. The NHIS benchmarks also included chronic pain, high-impact chronic pain, and headache or migraine bothering “a lot” in the past 3 months. We excluded refused, not ascertained, and don’t-know answers. We computed intervals by Taylor linearization with the published strata and primary sampling units. NHIS chronic pain (24.3%) and high-impact chronic pain (8.5%) matched published estimates.

#### Disclosure control

We suppressed counts of 1 to 20, rounded other counts up to a multiple of 10 (counts of 0 stayed 0), and, within each table, suppressed a further count wherever a suppressed count could otherwise be recovered from the table’s totals (complementary suppression). We computed conditional shares from the released counts. We released health-system results only as percentiles across sites and bootstrap results only as interval endpoints. All analyses of participant data ran inside the Researcher Workbench, and we exported only these aggregates.

#### Rounding sensitivity

The primary estimates (Table 2 in the main text) use the directly released counts for the primary analytic population, and their Wilson intervals treat these counts as exact. A released count $r > 0$ is compatible with any true count from $\max(21, r-9)$ to $r$. We therefore recomputed the shares and anchor-relative shares from 1,000 sets of counts, each count drawn uniformly and independently from its compatible values (redrawing pairs in which the coded count exceeded the number of self-reporters), and report the range of the resulting point estimates (eTable 6). For every condition except endometriosis and Lyme disease, the share and the anchor-relative share each varied by less than 0.01. For endometriosis, the share ranged from 0.407 to 0.416 (0.413 from the released counts) and the anchor-relative share from 0.487 to 0.497 (0.494). For Lyme disease, the condition with the smallest counts, the share ranged from 0.367 to 0.405 (0.394) and the anchor-relative share from 0.427 to 0.472 (0.459). For the conditions with lower shares, the estimate from the released counts lay near the upper end of its range, because rounding up adds about the same amount to the coded count as to the larger number of self-reporters, which raises their ratio.

**eTable 1. Eligibility and Analytic Populations.** Counts of participants, rounded up to a multiple of 10. Active EHR patients: All of Us participants born in 2004 or earlier with at least 1 EHR visit in each of 2 consecutive 12-month periods (July 2021 to June 2023). Record length was measured from the first EHR condition record (any condition) to the earliest survey completion date. Current care: a self-reported clinician diagnosis with ongoing visits or treatment (eTable 2). The current-care counts are the denominators of Table 2 in the main text. Bold: female predominant.

| Step | No. |
|---|---|
| Active adult EHR patients, any sex at birth | 262,790 |
| Excluded: sex at birth other or unknown | 2,300 |
| Female or male active EHR patients | 260,490 |
| Survey respondents (Personal and Family Health History or Personal Medical History) | 161,480 |
| Excluded: no survey date | 0 |
| Excluded: survey date but no EHR condition record | 2,400 |
| Excluded: first EHR condition record after the survey | 4,140 |
| Respondents with a first EHR condition record on or before the survey | 154,940 |
| Excluded: under 5 years of EHR condition records before the survey | 40,330 |
| Primary analytic population: 5 years or more of EHR condition records before the survey | 114,610 |
| *Primary analytic population reporting current care for the condition* | |
| Hypertension | 37,360 |
| Type 2 diabetes | 13,350 |
| Obesity | 16,080 |
| **Depression** | 28,130 |
| **Anxiety** | 21,450 |
| **Migraine** | 12,970 |
| **Fibromyalgia** | 5,350 |
| **Endometriosis** (women) | 1,500 |
| Lyme disease | 330 |
| **Chronic fatigue** | 4,880 |

**eTable 2. Conditions, ICD-10-CM Codes, and Survey Items.** A code was any ICD-10-CM code beginning with the listed prefix, recorded at any time in the participant's EHR condition table (for obesity, also the observation table). Ever-told self-report: any answer (including a skipped answer) to "About how old were you when you were first told you had [condition]?", which the surveys show only to participants who report the condition for themselves (eTable 3). For 9 conditions, participants do so by selecting themselves in "Including yourself, who in your family has had [condition]?" or in the earlier survey's condition list, and for Lyme disease, through a question about the respondent alone. Current self-report: ever told and "Yes" to "Are you still seeing a doctor or health care provider for [condition]?" or "Are you currently prescribed medications and/or receiving treatment for [condition]?" Concept IDs are All of Us survey question concepts. Long COVID and chronic pain have no survey item and were studied by comparison with national prevalence only. Bold: female predominant.

| Condition | ICD-10-CM codes | Ever told (concept ID) | Current care (concept IDs) |
|---|---|---|---|
| Hypertension | I10, I11, I12, I13, I15, I16 | 43530468 | 43530310, 1384437 |
| Type 2 diabetes | E11 | 43530492 | 43530335, 43528820 |
| Obesity | E66, Z68.3, Z68.4 | 43530542 | 43530384, 43528869 |
| **Depression** | F32, F33 | 43530516 | 43530358, 43528843 |
| **Anxiety** | F41 | 43530514 | 43530356, 43528841 |
| **Migraine** | G43 | 43530529 | 43530371, 43528856 |
| **Fibromyalgia** | M79.7 | 43530551 | 43530393, 43528878 |
| **Endometriosis** | N80 | 1384636 | 1384474, 1384624 |
| Lyme disease | A69.2 | 1384432 | 1384431, 1384628 |
| **Chronic fatigue** | G93.3, R53.82 | 43530523 | 43530365, 43528850 |
| **Long COVID** | U09.9 | – | – |
| Chronic pain | G89.2, G89.4 | – | – |

**eTable 3. Survey Routing and Item Nonresponse.** All survey respondents (all lengths of EHR record, and women only for endometriosis), counted once across the Personal and Family Health History and Personal Medical History surveys. Selected themselves: reported the condition for themselves (in the family-history question or the earlier survey's condition list, or, for Lyme disease, in a question about the respondent alone that every respondent was shown). First-told answer: any answer to the first-told question, the ever-told definition of the analysis. Skipped: the recorded answer was a skip. We count these participants as ever told. Care questions unanswered: a first-told answer but no answer to either current-care question. $\leq$20: a count of 1 to 20, suppressed. Suppressed: withheld so that a suppressed count cannot be recovered from the row. Survey version numbers were not populated in the data, so routing was checked by survey (eMethods).

| Condition | Selected themselves | First-told answer | Selected, no first-told answer | First-told answer, not selected | First-told skipped | Care questions unanswered | Current care |
|---|---|---|---|---|---|---|---|
| Hypertension | 53,400 | Suppressed | $\leq$20 | 0 | 200 | 0 | 49,550 |
| Type 2 diabetes | 18,470 | Suppressed | $\leq$20 | 0 | 80 | 0 | 17,660 |
| Obesity | 42,600 | Suppressed | $\leq$20 | 0 | 160 | 0 | 21,230 |
| **Depression** | 51,770 | Suppressed | $\leq$20 | 0 | 150 | 0 | 38,570 |
| **Anxiety** | 37,700 | 37,690 | $\leq$20 | $\leq$20 | 140 | 0 | 29,970 |
| **Migraine** | 32,380 | 32,380 | $\leq$20 | $\leq$20 | 80 | 0 | 17,490 |
| **Fibromyalgia** | 9,680 | 9,680 | $\leq$20 | $\leq$20 | 40 | 0 | 6,980 |
| **Endometriosis** (women) | 10,100 | Suppressed | $\leq$20 | 0 | 40 | 0 | 2,120 |
| Lyme disease | Suppressed | 4,460 | 0 | $\leq$20 | 60 | 0 | 430 |
| **Chronic fatigue** | 9,560 | Suppressed | $\leq$20 | 0 | 40 | 0 | 6,620 |

**eTable 4. Survey Occasions and Sensitivity Analysis Using the Earliest Survey.** Primary analytic population (5 years or more of EHR condition records before the survey). Of these participants, 13,240 of 114,610 (11.6%) answered on 2 or more survey occasions. A survey occasion is one questionnaire response. Answers on another occasion: participants in the current-care group (Table 2 in the main text) who, on another occasion, answered the condition's survey questions without reporting current care, given as No. (% of the current-care group). Earliest occasion: current care reported on the participant's earliest survey occasion, with a qualifying code at any date or dated no later than 1 year after that occasion. We computed proportions and 95% Wilson CIs from the released counts. Primary analysis: answers from any occasion and codes from any date (Table 2). Bold: female predominant.

| Condition | Answers on another occasion, No. (%) | Earliest occasion, No. | Coded, any date | Coded within 1 year | Primary analysis |
|---|---|---|---|---|---|
| Hypertension | 4,660 (12.5) | 33,790 | 0.84 (0.84–0.84) | 0.82 (0.82–0.83) | 0.85 |
| Type 2 diabetes | 1,690 (12.7) | 12,070 | 0.86 (0.86–0.87) | 0.85 (0.85–0.86) | 0.87 |
| Obesity | 2,000 (12.4) | 14,600 | 0.74 (0.74–0.75) | 0.71 (0.70–0.72) | 0.75 |
| **Depression** | 3,270 (11.6) | 25,770 | 0.67 (0.67–0.68) | 0.64 (0.63–0.64) | 0.68 |
| **Anxiety** | 2,560 (11.9) | 19,630 | 0.67 (0.66–0.68) | 0.63 (0.63–0.64) | 0.68 |
| **Migraine** | 1,560 (12.0) | 11,940 | 0.66 (0.66–0.67) | 0.63 (0.62–0.64) | 0.67 |
| **Fibromyalgia** | 640 (12.0) | 5,170 | 0.62 (0.61–0.64) | 0.60 (0.59–0.61) | 0.63 |
| **Endometriosis** (women) | 160 (10.7) | 1,450 | 0.41 (0.38–0.43) | 0.38 (0.35–0.40) | 0.41 |
| Lyme disease | 0 (0.0) | 310 | 0.39 (0.33–0.44) | 0.39 (0.33–0.44) | 0.39 |
| **Chronic fatigue** | 570 (11.7) | 4,750 | 0.24 (0.23–0.26) | 0.22 (0.21–0.23) | 0.24 |

**eTable 5. National Prevalence Benchmarks.** Weighted prevalence among US adults, with design-based 95% CIs. NHANES: examination weights, adults 18 years or older for hypertension and 20 or older otherwise, with pregnant women excluded for obesity. NHIS: Sample Adult weights. Female predominant: women-to-men ratio of 1.5 or more, or a condition defined only in women (endometriosis). We classified fibromyalgia and Lyme disease, which have no national item, from self-reported diagnosis among All of Us respondents (women-to-men ratios 5.56 and 0.86).

| Measure | Survey | Women | Men | Ratio |
|---|---|---|---|---|
| Hypertension, measured or treated | NHANES | 0.446 (0.419–0.473) | 0.511 (0.489–0.534) | 0.87 |
| Hypertension, ever told | NHIS | 0.310 (0.301–0.318) | 0.336 (0.327–0.346) | 0.92 |
| Diabetes (any type), diagnosed | NHANES | 0.111 (0.092–0.130) | 0.124 (0.110–0.137) | 0.90 |
| Type 2 diabetes, ever told | NHIS | 0.082 (0.077–0.087) | 0.083 (0.078–0.088) | 0.99 |
| Obesity, measured | NHANES | 0.413 (0.370–0.456) | 0.392 (0.354–0.430) | 1.05 |
| Depression, ever told | NHIS | 0.238 (0.230–0.247) | 0.137 (0.130–0.144) | 1.74 |
| Anxiety, ever told | NHIS | 0.238 (0.230–0.246) | 0.129 (0.122–0.136) | 1.85 |
| Headache or migraine bothered a lot (past 3 mo) | NHIS | 0.078 (0.073–0.084) | 0.032 (0.028–0.035) | 2.49 |
| ME/CFS, current | NHIS | 0.015 (0.013–0.018) | 0.006 (0.005–0.008) | 2.38 |
| Long COVID, ever | NHIS | 0.102 (0.096–0.108) | 0.064 (0.059–0.069) | 1.60 |
| Long COVID, current | NHIS | 0.046 (0.042–0.050) | 0.025 (0.022–0.028) | 1.84 |
| High-impact chronic pain | NHIS | 0.096 (0.091–0.101) | 0.073 (0.068–0.078) | 1.32 |

**eTable 6. Sensitivity of the Primary Estimates to Rounding of Released Counts.** Current self-reporters with 5 or more years of EHR condition records before the survey (Table 2 in the main text, columns All and Relative to anchors). A released count $r > 0$ (rounded up to a multiple of 10, with counts of 1 to 20 suppressed) is compatible with any true count from $\max(21, r - 9)$ to $r$. In each of 1,000 draws, every count entering an estimate (coded and current self-reporters, and the counts behind the anchor mean) was drawn uniformly and independently from its compatible values, and pairs in which the coded count exceeded the number of self-reporters were redrawn. Estimate: from the released counts (as in Table 2). Range: minimum to maximum of the 1,000 point estimates. Relative to anchors: the share divided by the mean share for hypertension and type 2 diabetes (for endometriosis, the female anchors). We show the anchors relative to their own mean. Bold: female predominant.

| Condition | Share coded, estimate (range) | Relative to anchors, estimate (range) |
|---|---|---|
| Hypertension | 0.846 (0.846–0.846) | 0.986 (0.985–0.986) |
| Type 2 diabetes | 0.870 (0.870–0.871) | 1.014 (1.014–1.015) |
| Obesity | 0.753 (0.753–0.754) | 0.877 (0.877–0.878) |
| **Depression** | 0.678 (0.678–0.678) | 0.790 (0.789–0.790) |
| **Anxiety** | 0.679 (0.679–0.680) | 0.791 (0.791–0.792) |
| **Migraine** | 0.672 (0.672–0.673) | 0.783 (0.782–0.784) |
| **Fibromyalgia** | 0.626 (0.624–0.627) | 0.730 (0.728–0.731) |
| **Endometriosis** | 0.413 (0.407–0.416) | 0.494 (0.487–0.497) |
| Lyme disease | 0.394 (0.367–0.405) | 0.459 (0.427–0.472) |
| **Chronic fatigue** | 0.244 (0.242–0.244) | 0.284 (0.282–0.285) |

**eTable 7. Interval Estimates for the Share Relative to Anchors.** Primary analytic population (Table 2 in the main text). Participant bootstrap: 2,000 replicates with Poisson(1) weights per participant, keeping each participant's membership in every condition and both anchors, computed on unrounded data within the Researcher Workbench, with percentile 95% CIs (used in the main text). Site-cluster bootstrap: the same with health systems (assigned EHR sites) resampled, so that it reflects variation between health systems. Fixed-anchor approximation: the Wilson limits of the share divided by the anchor mean, treating the mean as fixed. Relative share: from the released counts. Endometriosis: relative to the female anchors. Bold: female predominant.

| Condition | Relative share | Participant bootstrap | Site-cluster bootstrap | Fixed-anchor approximation |
|---|---|---|---|---|
| Obesity | 0.88 | 0.87–0.89 | 0.83–0.92 | 0.87–0.89 |
| **Depression** | 0.79 | 0.78–0.80 | 0.72–0.84 | 0.78–0.80 |
| **Anxiety** | 0.79 | 0.78–0.80 | 0.71–0.86 | 0.78–0.80 |
| **Migraine** | 0.78 | 0.77–0.79 | 0.71–0.84 | 0.77–0.79 |
| **Fibromyalgia** | 0.73 | 0.71–0.75 | 0.67–0.78 | 0.71–0.74 |
| **Endometriosis** (women) | 0.49 | 0.46–0.52 | 0.42–0.55 | 0.46–0.52 |
| Lyme disease | 0.46 | 0.39–0.51 | 0.29–0.54 | 0.40–0.52 |
| **Chronic fatigue** | 0.28 | 0.27–0.30 | 0.24–0.33 | 0.27–0.30 |

**eTable 8. Share Coded Among Self-Reporters, All Lengths of EHR Record, by Sex.** Survey respondents among active adult EHR patients (104,250 women and 57,230 men, rounded). Proportions with 95% Wilson CIs from counts rounded up to a multiple of 10. NA: not applicable (endometriosis among men). Bold: female predominant.

| Condition | Women, ever | Men, ever | Women, current | Men, current |
|---|---|---|---|---|
| Hypertension | 0.78 (0.78–0.79) | 0.82 (0.82–0.83) | 0.80 (0.80–0.81) | 0.84 (0.83–0.84) |
| Type 2 diabetes | 0.81 (0.80–0.82) | 0.88 (0.87–0.88) | 0.82 (0.81–0.83) | 0.88 (0.87–0.88) |
| Obesity | 0.66 (0.65–0.66) | 0.68 (0.67–0.69) | 0.71 (0.70–0.72) | 0.74 (0.72–0.75) |
| **Depression** | 0.57 (0.56–0.57) | 0.59 (0.58–0.60) | 0.63 (0.62–0.63) | 0.67 (0.66–0.68) |
| **Anxiety** | 0.61 (0.60–0.61) | 0.61 (0.60–0.62) | 0.64 (0.63–0.65) | 0.66 (0.65–0.67) |
| **Migraine** | 0.49 (0.48–0.50) | 0.41 (0.39–0.42) | 0.65 (0.64–0.65) | 0.61 (0.59–0.63) |
| **Fibromyalgia** | 0.55 (0.54–0.56) | 0.39 (0.36–0.42) | 0.62 (0.60–0.63) | 0.45 (0.41–0.49) |
| **Endometriosis** | 0.17 (0.17–0.18) | NA | 0.38 (0.36–0.40) | NA |
| Lyme disease | 0.19 (0.18–0.21) | 0.20 (0.18–0.21) | 0.35 (0.30–0.41) | 0.38 (0.31–0.47) |
| **Chronic fatigue** | 0.21 (0.20–0.21) | 0.19 (0.17–0.20) | 0.22 (0.21–0.24) | 0.21 (0.19–0.23) |

**eTable 9. Coverage of the Code Search.** Current self-reporters in the primary analytic population (Table 2 in the main text), classified by the first of these that applies: a qualifying ICD-10-CM code (the primary definition), only an ICD-9-CM code for the condition, only a code from another source vocabulary (not ICD-10-CM and not survey) whose standard concept is one to which a qualifying ICD-10-CM code maps, or none of these. No participant had a qualifying ICD-10-CM code only in the observation table (for obesity, the primary definition already includes that table). Any record: share with a record in any of the 3 categories. No. (%). ≤20: a count of 1 to 20, suppressed. Suppressed: withheld so that a suppressed count cannot be recovered from the row. Bold: female predominant. ICD-9-CM codes searched (number of mapped standard concepts): 401–405 for hypertension (16), 250.x0 and 250.x2 for type 2 diabetes (21), 278.0, V85.3, and V85.4 for obesity (8), 296.2, 296.3, and 311 for depression (18), 300.0 for anxiety (3), 346 for migraine (18), 729.1 for fibromyalgia (1), 617 for endometriosis (27), 088.81 for Lyme disease (3), and 780.71 for chronic fatigue (none for R53.82) (2 + 1).

| Condition | No. | ICD-10-CM (primary) | ICD-9-CM only | Other vocabulary only | None | Any record, % |
|---|---|---|---|---|---|---|
| Hypertension | 37,360 | 31,610 (84.6) | 3,110 (8.3) | 1,000 (2.7) | 1,650 (4.4) | 95.6 |
| Type 2 diabetes | 13,350 | 11,620 (87.0) | 1,070 (8.0) | 350 (2.6) | 330 (2.5) | 97.5 |
| Obesity | 16,080 | 12,110 (75.3) | 1,820 (11.3) | 720 (4.5) | 1,430 (8.9) | 91.1 |
| **Depression** | 28,130 | 19,070 (67.8) | 3,090 (11.0) | 1,280 (4.6) | 4,710 (16.7) | 83.3 |
| **Anxiety** | 21,450 | 14,570 (67.9) | 2,460 (11.5) | 710 (3.3) | 3,730 (17.4) | 82.6 |
| **Migraine** | 12,970 | 8,720 (67.2) | 1,550 (12.0) | 670 (5.2) | 2,040 (15.7) | 84.3 |
| **Fibromyalgia** | 5,350 | 3,350 (62.6) | 840 (15.7) | 210 (3.9) | 970 (18.1) | 81.9 |
| **Endometriosis** (women) | 1,500 | 620 (41.3) | 180 (12.0) | 50 (3.3) | 670 (44.7) | 55.3 |
| Lyme disease | 330 | 130 (39.4) | Suppressed | ≤20 | 160 (48.5) | 51.5 |
| **Chronic fatigue** | 4,880 | 1,190 (24.4) | 90 (1.8) | 110 (2.3) | 3,500 (71.7) | 28.3 |

**eTable 10. Share Coded Among Current Self-Reporters, by Years of EHR Condition Records Before the Survey.** Women and men pooled (endometriosis, women). After survey: the first EHR condition record follows the survey date. Proportions from counts rounded up to a multiple of 10. S: suppressed by disclosure control. Bold: female predominant.

| Condition | After survey | $<2$ | 2–4 | 5–9 | $\geq 10$ |
|---|---|---|---|---|---|
| Hypertension | 0.69 | 0.75 | 0.78 | 0.81 | 0.86 |
| Type 2 diabetes | S | S | 0.81 | 0.84 | 0.89 |
| Obesity | S | 0.59 | 0.67 | 0.71 | 0.78 |
| **Depression** | 0.36 | 0.54 | 0.59 | 0.64 | 0.70 |
| **Anxiety** | 0.38 | 0.55 | 0.61 | 0.65 | 0.70 |
| **Migraine** | 0.36 | 0.55 | 0.60 | 0.64 | 0.69 |
| **Fibromyalgia** | S | S | 0.60 | 0.60 | 0.64 |
| **Endometriosis** | S | 0.33 | 0.36 | 0.41 | 0.41 |
| Lyme disease | S | S | S | 0.40 | 0.39 |
| **Chronic fatigue** | S | S | 0.20 | 0.23 | 0.25 |
| Anchor mean | S | S | 0.80 | 0.83 | 0.88 |
| Female anchor mean | 0.72 | 0.78 | 0.77 | 0.81 | 0.86 |

Anchor mean: mean of the hypertension and type 2 diabetes shares in the group (S when either is suppressed). Female anchor mean: the same among women, the reference for endometriosis.

**eTable 11. Variation Across Health Systems.** Survey respondents, all lengths of EHR record. Each patient was assigned to the EHR site contributing most of their visits from July 2021 to June 2023 (participant-mediated EHR excluded). A site contributes a condition's share when it has at least 50 current self-reporters of the condition (and of each anchor, for the relative share), so that each site's share has a standard error of at most 0.07. Not released: fewer than 5 such sites. Values are the median (10th–90th percentile) across sites, with no site identified. Endometriosis uses the female anchors. Bold: female predominant.

| Condition | Sites | Share coded | Relative to site's anchors |
|---|---|---|---|
| Hypertension | 51 | 0.92 (0.30–0.97) | 0.99 (0.96–1.01) |
| Type 2 diabetes | 44 | 0.95 (0.32–0.98) | 1.01 (0.99–1.04) |
| Obesity | 40 | 0.83 (0.18–0.91) | 0.90 (0.70–0.96) |
| **Depression** | 46 | 0.72 (0.11–0.84) | 0.79 (0.55–0.89) |
| **Anxiety** | 45 | 0.75 (0.10–0.85) | 0.81 (0.58–0.91) |
| **Migraine** | 39 | 0.74 (0.22–0.82) | 0.79 (0.56–0.87) |
| **Fibromyalgia** | 31 | 0.69 (0.29–0.78) | 0.76 (0.65–0.85) |
| **Endometriosis** | 12 | 0.47 (0.07–0.64) | 0.49 (0.18–0.69) |
| Lyme disease | fewer than 5 sites | Not released | |
| **Chronic fatigue** | 30 | 0.25 (0.07–0.38) | 0.29 (0.13–0.43) |

**eTable 12. Women-to-Men Ratios of the Share Coded: Crude, Standardized, and Within Health System.** Primary analytic population, current self-reporters. Crude: Table 2 in the main text. Standardized: each sex's share directly standardized to the pooled distribution of current self-reporters of both sexes over age band (5 bands) and record length (5 to 9 years, 10 years or more). Within site: Mantel-Haenszel ratio over strata of assigned EHR site, age band, and record length. Sites: sites contributing strata with both sexes. 95% CIs from the participant-level bootstrap (eMethods). Bold: female predominant. Endometriosis was analyzed among women only.

| Condition | Crude ratio (95% CI) | Standardized share, women | Standardized share, men | Standardized ratio (95% CI) | Within-site ratio (95% CI) | Sites |
|---|---|---|---|---|---|---|
| Hypertension | 0.96 (0.95–0.97) | 0.84 | 0.86 | 0.98 (0.97–0.98) | 0.99 (0.98–1.00) | 56 |
| Type 2 diabetes | 0.94 (0.92–0.95) | 0.85 | 0.89 | 0.96 (0.94–0.97) | 0.98 (0.97–0.99) | 54 |
| Obesity | 0.95 (0.93–0.97) | 0.75 | 0.77 | 0.97 (0.95–0.99) | 1.00 (0.98–1.02) | 52 |
| **Depression** | 0.93 (0.92–0.95) | 0.67 | 0.70 | 0.95 (0.94–0.97) | 1.02 (1.00–1.04) | 55 |
| **Anxiety** | 0.96 (0.94–0.98) | 0.68 | 0.70 | 0.97 (0.95–0.99) | 1.01 (0.99–1.03) | 53 |
| **Migraine** | 1.06 (1.03–1.10) | 0.68 | 0.63 | 1.07 (1.03–1.12) | 1.11 (1.07–1.15) | 46 |
| **Fibromyalgia** | 1.34 (1.24–1.51) | 0.64 | 0.49 | 1.30 (1.19–1.44) | 1.41 (1.29–1.57) | 41 |
| Lyme disease | 0.98 (0.73–1.32) | 0.38 | 0.38 | 1.01 (0.72–1.39) | 0.81 (0.57–1.13) | 12 |
| **Chronic fatigue** | 1.09 (0.95–1.23) | 0.25 | 0.22 | 1.11 (0.96–1.29) | 1.10 (0.95–1.28) | 40 |

**eTable 13. Share Coded Among Current Self-Reporters, by Race and Ethnicity.** Women and men pooled (endometriosis, women), all lengths of EHR record. NH, non-Hispanic. Proportions from counts rounded up to a multiple of 10. S: suppressed by disclosure control. Bold: female predominant.

| Condition | Hispanic | NH Asian | NH Black | NH White | NH other | Unknown |
|---|---|---|---|---|---|---|
| **A.** Share coded | | | | | | |
| Hypertension | 0.84 | 0.78 | 0.84 | 0.81 | 0.78 | 0.86 |
| Type 2 diabetes | 0.87 | S | 0.86 | 0.84 | 0.82 | S |
| Obesity | 0.74 | 0.64 | 0.78 | 0.71 | 0.68 | 0.76 |
| **Depression** | 0.65 | 0.57 | 0.67 | 0.64 | 0.62 | 0.66 |
| **Anxiety** | 0.65 | 0.58 | 0.63 | 0.65 | 0.60 | 0.67 |
| **Migraine** | 0.62 | 0.59 | 0.60 | 0.65 | 0.59 | 0.68 |
| **Fibromyalgia** | 0.65 | S | 0.63 | 0.59 | 0.56 | S |
| **Endometriosis** | 0.41 | S | 0.33 | 0.38 | 0.44 | S |
| Lyme disease | S | S | S | 0.38 | S | S |
| **Chronic fatigue** | 0.16 | S | S | 0.24 | 0.20 | S |
| Anchor mean | 0.86 | S | 0.85 | 0.83 | 0.80 | S |
| Female anchor mean | 0.84 | 0.79 | 0.84 | 0.80 | 0.77 | 0.88 |
| **B.** Share relative to anchors (95% CI) | | | | | | |
| Obesity | 0.87 (0.84–0.89) | S | 0.92 (0.90–0.94) | 0.85 (0.85–0.86) | 0.85 (0.82–0.89) | S |
| **Depression** | 0.76 (0.74–0.78) | S | 0.79 (0.77–0.80) | 0.77 (0.76–0.78) | 0.77 (0.75–0.80) | S |
| **Anxiety** | 0.76 (0.74–0.78) | S | 0.74 (0.72–0.77) | 0.78 (0.78–0.79) | 0.75 (0.73–0.79) | S |
| **Migraine** | 0.72 (0.69–0.75) | S | 0.71 (0.68–0.74) | 0.79 (0.78–0.80) | 0.74 (0.70–0.78) | S |
| **Fibromyalgia** | 0.76 (0.71–0.79) | S | 0.74 (0.69–0.78) | 0.72 (0.70–0.73) | 0.70 (0.65–0.76) | S |
| **Endometriosis** | 0.48 (0.41–0.56) | S | 0.40 (0.28–0.45) | 0.48 (0.45–0.51) | 0.57 (0.41–0.61) | S |
| Lyme disease | S | S | S | 0.46 (0.39–0.51) | S | S |
| **Chronic fatigue** | 0.19 (0.16–0.23) | S | S | 0.29 (0.28–0.30) | 0.25 (0.20–0.29) | S |

Anchor mean: mean of the hypertension and type 2 diabetes shares in the group (S when either is suppressed). Female anchor mean: the same among women, the reference for endometriosis. Panel B: the share divided by the group's anchor mean (endometriosis: female anchor mean), with participant-level bootstrap 95% CIs. S also marks estimates whose bootstrap interval was suppressed.

**eTable 14. Share Coded Among Current Self-Reporters, by Age Band.** Women and men pooled (endometriosis, women), all lengths of EHR record. Age in years is 2023 minus birth year. Proportions from counts rounded up to a multiple of 10. S: suppressed by disclosure control. Bold: female predominant.

| Condition | 18–39 | 40–49 | 50–59 | 60–69 | ≥70 |
|---|---|---|---|---|---|
| **A.** Share coded | | | | | |
| Hypertension | 0.64 | 0.72 | 0.79 | 0.82 | 0.86 |
| Type 2 diabetes | 0.73 | 0.75 | 0.81 | 0.86 | 0.89 |
| Obesity | 0.65 | 0.69 | 0.72 | 0.74 | 0.75 |
| **Depression** | 0.57 | 0.60 | 0.64 | 0.66 | 0.72 |
| **Anxiety** | 0.62 | 0.64 | 0.64 | 0.66 | 0.69 |
| **Migraine** | 0.62 | 0.63 | 0.64 | 0.66 | 0.66 |
| **Fibromyalgia** | 0.55 | 0.60 | 0.60 | 0.62 | 0.62 |
| **Endometriosis** | 0.44 | 0.44 | 0.32 | 0.20 | S |
| Lyme disease | S | S | 0.50 | 0.42 | 0.33 |
| **Chronic fatigue** | 0.22 | 0.22 | 0.21 | 0.24 | 0.27 |
| Anchor mean | 0.69 | 0.74 | 0.80 | 0.84 | 0.88 |
| Female anchor mean | 0.68 | 0.73 | 0.78 | 0.83 | 0.86 |
| **B.** Share relative to anchors (95% CI) | | | | | |
| Obesity | 0.95 (0.92–0.99) | 0.93 (0.91–0.95) | 0.90 (0.89–0.92) | 0.88 (0.86–0.89) | 0.86 (0.85–0.87) |
| **Depression** | 0.84 (0.81–0.87) | 0.81 (0.79–0.83) | 0.81 (0.79–0.82) | 0.79 (0.78–0.80) | 0.82 (0.81–0.84) |
| **Anxiety** | 0.91 (0.88–0.94) | 0.86 (0.84–0.88) | 0.80 (0.79–0.82) | 0.79 (0.77–0.80) | 0.79 (0.77–0.81) |
| **Migraine** | 0.90 (0.87–0.94) | 0.86 (0.84–0.89) | 0.80 (0.78–0.82) | 0.79 (0.77–0.81) | 0.76 (0.74–0.78) |
| **Fibromyalgia** | 0.79 (0.74–0.85) | 0.81 (0.78–0.85) | 0.76 (0.73–0.78) | 0.74 (0.71–0.76) | 0.71 (0.68–0.74) |
| **Endometriosis** | 0.65 (0.59–0.70) | 0.61 (0.55–0.66) | 0.42 (0.36–0.49) | 0.24 (0.12–0.26) | S |
| Lyme disease | S | S | 0.63 (0.41–0.69) | 0.49 (0.38–0.60) | 0.38 (0.30–0.50) |
| **Chronic fatigue** | 0.31 (0.28–0.34) | 0.29 (0.26–0.32) | 0.26 (0.23–0.28) | 0.28 (0.25–0.31) | 0.30 (0.26–0.33) |

Anchor mean: mean of the hypertension and type 2 diabetes shares in the group (S when either is suppressed). Female anchor mean: the same among women, the reference for endometriosis. Panel B: the share divided by the group's anchor mean (endometriosis: female anchor mean), with participant-level bootstrap 95% CIs. S also marks estimates whose bootstrap interval was suppressed.

**eTable 15. Coded Share Standardized to US Adults Divided by National Prevalence.** Coded share: any code among all active adult EHR patients of that sex, directly standardized to the US adult age (or age by race and ethnicity) distribution from NHANES 2021-2023 interview weights. Ages: adults 18 years or older, or, for the NHANES obesity and diagnosed-diabetes benchmarks (adults 20 or older), active patients aged 20 or older standardized to US adults aged 20 or older. For obesity among women, we excluded women with a pregnancy code from July 2021 to June 2023, as NHANES excludes pregnant women. Diabetes, any type: codes E08 to E11 and E13, compared with NHANES diagnosed diabetes of any type. Comparisons of chronic pain codes with high-impact chronic pain, migraine codes with headache or migraine bothering the respondent a lot, and G93.3 codes with current ME/CFS involve nonequivalent definitions and are exploratory. Bold: female predominant.

| Condition | Benchmark | Ages | Standard | Sex | Coded | National | Ratio (95% CI) |
|---|---|---|---|---|---|---|---|
| Hypertension | NHANES measured | 18+ | Age | F | 0.345 | 0.446 | 0.77 (0.73–0.82) |
| Hypertension | NHANES measured | 18+ | Age, race | F | 0.335 | 0.446 | 0.75 (0.71–0.80) |
| Hypertension | NHANES measured | 18+ | Age | M | 0.411 | 0.511 | 0.80 (0.77–0.84) |
| Hypertension | NHANES measured | 18+ | Age, race | M | 0.406 | 0.511 | 0.79 (0.76–0.83) |
| Hypertension | NHIS ever told | 18+ | Age | F | 0.345 | 0.310 | 1.12 (1.08–1.15) |
| Hypertension | NHIS ever told | 18+ | Age, race | F | 0.335 | 0.310 | 1.08 (1.05–1.11) |
| Hypertension | NHIS ever told | 18+ | Age | M | 0.411 | 0.336 | 1.22 (1.19–1.26) |
| Hypertension | NHIS ever told | 18+ | Age, race | M | 0.406 | 0.336 | 1.21 (1.17–1.24) |
| Type 2 diabetes | NHIS ever told | 18+ | Age | F | 0.162 | 0.082 | 1.97 (1.85–2.10) |
| Type 2 diabetes | NHIS ever told | 18+ | Age, race | F | 0.154 | 0.082 | 1.88 (1.77–2.00) |
| Type 2 diabetes | NHIS ever told | 18+ | Age | M | 0.191 | 0.083 | 2.31 (2.16–2.46) |
| Type 2 diabetes | NHIS ever told | 18+ | Age, race | M | 0.189 | 0.083 | 2.28 (2.13–2.43) |
| Diabetes, any type | NHANES diagnosed | 20+ | Age | F | 0.169 | 0.111 | 1.52 (1.28–1.80) |
| Diabetes, any type | NHANES diagnosed | 20+ | Age, race | F | 0.161 | 0.111 | 1.46 (1.23–1.72) |
| Diabetes, any type | NHANES diagnosed | 20+ | Age | M | 0.201 | 0.124 | 1.62 (1.46–1.81) |
| Diabetes, any type | NHANES diagnosed | 20+ | Age, race | M | 0.198 | 0.124 | 1.60 (1.44–1.78) |
| Obesity | NHANES measured | 20+ | Age | F | 0.368 | 0.413 | 0.89 (0.80–0.99) |
| Obesity | NHANES measured | 20+ | Age, race | F | 0.356 | 0.413 | 0.86 (0.78–0.96) |
| Obesity | NHANES measured | 20+ | Age | M | 0.320 | 0.392 | 0.82 (0.74–0.90) |
| Obesity | NHANES measured | 20+ | Age, race | M | 0.317 | 0.392 | 0.81 (0.73–0.89) |
| **Depression** | NHIS ever told | 18+ | Age | F | 0.340 | 0.238 | 1.43 (1.38–1.48) |
| **Depression** | NHIS ever told | 18+ | Age, race | F | 0.334 | 0.238 | 1.40 (1.35–1.45) |
| **Depression** | NHIS ever told | 18+ | Age | M | 0.285 | 0.137 | 2.08 (1.97–2.20) |
| **Depression** | NHIS ever told | 18+ | Age, race | M | 0.281 | 0.137 | 2.06 (1.95–2.17) |
| **Anxiety** | NHIS ever told | 18+ | Age | F | 0.385 | 0.238 | 1.62 (1.56–1.68) |
| **Anxiety** | NHIS ever told | 18+ | Age, race | F | 0.381 | 0.238 | 1.60 (1.54–1.66) |
| **Anxiety** | NHIS ever told | 18+ | Age | M | 0.304 | 0.129 | 2.36 (2.23–2.49) |
| **Anxiety** | NHIS ever told | 18+ | Age, race | M | 0.304 | 0.129 | 2.36 (2.23–2.49) |
| **Migraine** | NHIS headache a lot | 18+ | Age | F | 0.204 | 0.078 | 2.60 (2.43–2.78) |
| **Migraine** | NHIS headache a lot | 18+ | Age, race | F | 0.201 | 0.078 | 2.57 (2.40–2.74) |
| **Migraine** | NHIS headache a lot | 18+ | Age | M | 0.087 | 0.032 | 2.77 (2.46–3.12) |
| **Migraine** | NHIS headache a lot | 18+ | Age, race | M | 0.087 | 0.032 | 2.77 (2.45–3.12) |
| **ME/CFS and postviral fatigue** | NHIS current | 18+ | Age | F | 0.009 | 0.015 | 0.57 (0.49–0.67) |
| **ME/CFS and postviral fatigue** | NHIS current | 18+ | Age, race | F | 0.009 | 0.015 | 0.57 (0.49–0.67) |
| **ME/CFS and postviral fatigue** | NHIS current | 18+ | Age | M | 0.005 | 0.006 | 0.72 (0.55–0.94) |
| **ME/CFS and postviral fatigue** | NHIS current | 18+ | Age, race | M | 0.005 | 0.006 | 0.72 (0.55–0.94) |
| **Long COVID** | NHIS current | 18+ | Age | F | 0.011 | 0.046 | 0.24 (0.22–0.27) |
| **Long COVID** | NHIS current | 18+ | Age, race | F | 0.011 | 0.046 | 0.24 (0.22–0.26) |
| **Long COVID** | NHIS current | 18+ | Age | M | 0.008 | 0.025 | 0.31 (0.27–0.36) |
| **Long COVID** | NHIS current | 18+ | Age, race | M | 0.008 | 0.025 | 0.31 (0.27–0.36) |
| **Long COVID** | NHIS ever | 18+ | Age | F | 0.011 | 0.102 | 0.11 (0.10–0.12) |
| **Long COVID** | NHIS ever | 18+ | Age, race | F | 0.011 | 0.102 | 0.11 (0.10–0.12) |
| **Long COVID** | NHIS ever | 18+ | Age | M | 0.008 | 0.064 | 0.12 (0.11–0.14) |
| **Long COVID** | NHIS ever | 18+ | Age, race | M | 0.008 | 0.064 | 0.12 (0.11–0.14) |
| Chronic pain | NHIS high-impact | 18+ | Age | F | 0.354 | 0.096 | 3.69 (3.49–3.90) |
| Chronic pain | NHIS high-impact | 18+ | Age, race | F | 0.348 | 0.096 | 3.62 (3.43–3.83) |
| Chronic pain | NHIS high-impact | 18+ | Age | M | 0.302 | 0.073 | 4.15 (3.86–4.46) |
| Chronic pain | NHIS high-impact | 18+ | Age, race | M | 0.298 | 0.073 | 4.10 (3.81–4.41) |

Sex: F indicates women and M indicates men. Standard: age, or age and race and ethnicity.
